# Neurocognitive and resting-state functional MRI changes in patients with diffuse gliomas after chemoradiotherapy

**DOI:** 10.1101/2024.09.25.24314312

**Authors:** Zhihua Liu, Timothy J. Mitchell, Chongliang Luo, Ki Yun Park, Joshua S. Shimony, Robert Fucetola, Eric C. Leuthardt, Stephanie M. Perkins, Abraham Z. Snyder, Tong Zhu, Jiayi Huang

**Author notes:** Equal Contribution (Zhihua Liu, Timothy J. Mitchell). **Corresponding Authors:** Jiayi Huang, MD, MSCI, Department of Radiation Oncology, School of Medicine, Washington University in St. Louis, 4921 Parkview Place, Campus Box 8224, St. Louis, MO 63110.

## Abstract

**Background:** This prospective observational study employed resting-state functional MRI (rs-fMRI) to investigate network-level disturbances associated with neurocognitive function (NCF) changes in patients with gliomas following radiation therapy (RT).

**Methods:** Adult patients with either IDH-wildtype or IDH-mutant gliomas underwent computerized NCF testing and rs-fMRI before and 6 months after RT. NCF changes were quantified by the percent change in age-normalized composite scores from baseline (ΔNCF_comp_). rs-fMRI data underwent seed-based functional connectivity (FC) analysis. Whole-brain connectivity regression analysis assessed the association between network FC changes and NCF changes, using a split-sample approach with a 26-patient training set and a 6-patient validation set, iterated 200 times. Permutation tests evaluated the significance of network selection.

**Results:** Between September 2020 and December 2023, 43 patients were enrolled, with 32 completing both initial and follow-up evaluations. The mean ΔNCF_comp_ was 2.9% (SD: 13.7%), with 38% experiencing a decline. Intra-hemispheric FC remained similar between ipsilateral and contralateral hemispheres for most patients at both time points. FC changes accounted for a moderate amount of variance in NCF changes (mean R^2^: 0.301, SD: 0.249), with intra-network FC of the Parietal Memory Network (PMN-PMN, *P*=0.001) and inter-network FC between the PMN and the Visual Network (PMN-VN, *P*=0.002) as the most significant factors. Similar findings were obtained by sensitivity analyses using only the FC data from the hemisphere contralateral to the tumor.

**Conclusions:** Post-RT rs-fMRI changes significantly predicted NCF decline, highlighting rs-fMRI as a promising imaging biomarker for neurocognitive decline after RT.

## INTRODUCTION

Diffuse gliomas comprise approximately 80% of all primary malignant brain tumors in adults. There were an estimated 141,446 individuals living with a glioma diagnosis in 2019, or approximately 11% of the total population diagnosed with brain tumors.^1^ These tumors are broadly categorized by mutations in the isocitrate dehydrogenase (IDH) gene and are treated with surgery followed by chemoradiotherapy.^2–4^ The IDH-wildtype glioma consists mostly of the aggressive glioblastoma (GBM, WHO grade 4), which has a median overall survival (OS) of approximately 16 months after chemoradiotherapy.^5^ The more favorable IDH-mutant gliomas can be characterized as either astrocytoma or oligodendroglioma, with a median OS of approximately 7 and 14 years after chemoradiotherapy, respectively.^2,6,7^ Neurocognitive function (NCF) decline after chemoradiotherapy represents a major treatment complication in brain tumor survivors and is associated with reduced quality of life.^8,9^ Since IDH-mutant gliomas typically affect a younger population and have longer survival, NCF impairment may cause greater societal and economic loss.^10^ A prior large, randomized study of patients with GBM showed that approximately 36% of the progression-free patients developed significant NCF decline 6 months after RT, mostly involving impairment of episodic memory, executive function, and processing speed.^11^ However, it is currently unknown what regions of the brain are most vulnerable to radiation injury and contribute the most to NCF decline following partial-brain RT for glioma.

Resting-state functional magnetic resonance imaging (rs-fMRI) is an advanced imaging method that identifies the spatiotemporal distribution of intrinsic functional networks within the brain.^12^ rs-fMRI measures the temporal correlation of spontaneous low-frequency fluctuations (<0.1 Hz) in the blood-oxygen-level-dependent (BOLD) signal in the task-free (“resting”) state. Regions of the brain exhibiting strong correlations are widely known as resting-state networks (RSNs).^13,14^ The network assignment mapped by the individual rs-fMRI RSN topography generally matches functional anatomy observed by other methods, including task-based functional MRI and intra-operative electrocortical stimulation mapping.^15–17^ rs-fMRI has been widely used in system neuroscience research in Alzheimer’s dementia, stroke, and aging.^18–20^ The advantage of using rs-fMRI over task-based fMRI is that one can simultaneously evaluate multiple networks in a single scan without having to design a specific task targeting a selected cognitive process.

We designed a prospective, longitudinal study to conduct rs-fMRI, NCF testing, and patient-reported outcomes (PROs) assessments on patients with gliomas before and after RT. In this first analysis, we correlated changes of NCF with changes of rs-fMRI and PROs. The primary aim of the current analysis is to assess whether changes of rs-fMRI can predict NCF decline after RT and to identify specific dominant network-level disruptions as candidate imaging biomarkers.

## METHODS

### Study Design

This prospective, single-institutional, observational study longitudinally evaluates patients with diffuse gliomas before and after standard fractionated RT. The study is designed to assess rs-fMRI, NCF, and patient-reported QOL at baseline (before RT), 6 months, 2 years, and 5 years after RT (**Figure 1A**). This paper presents initial findings from the baseline and 6-month post-RT assessments.

**Fig. 1.**
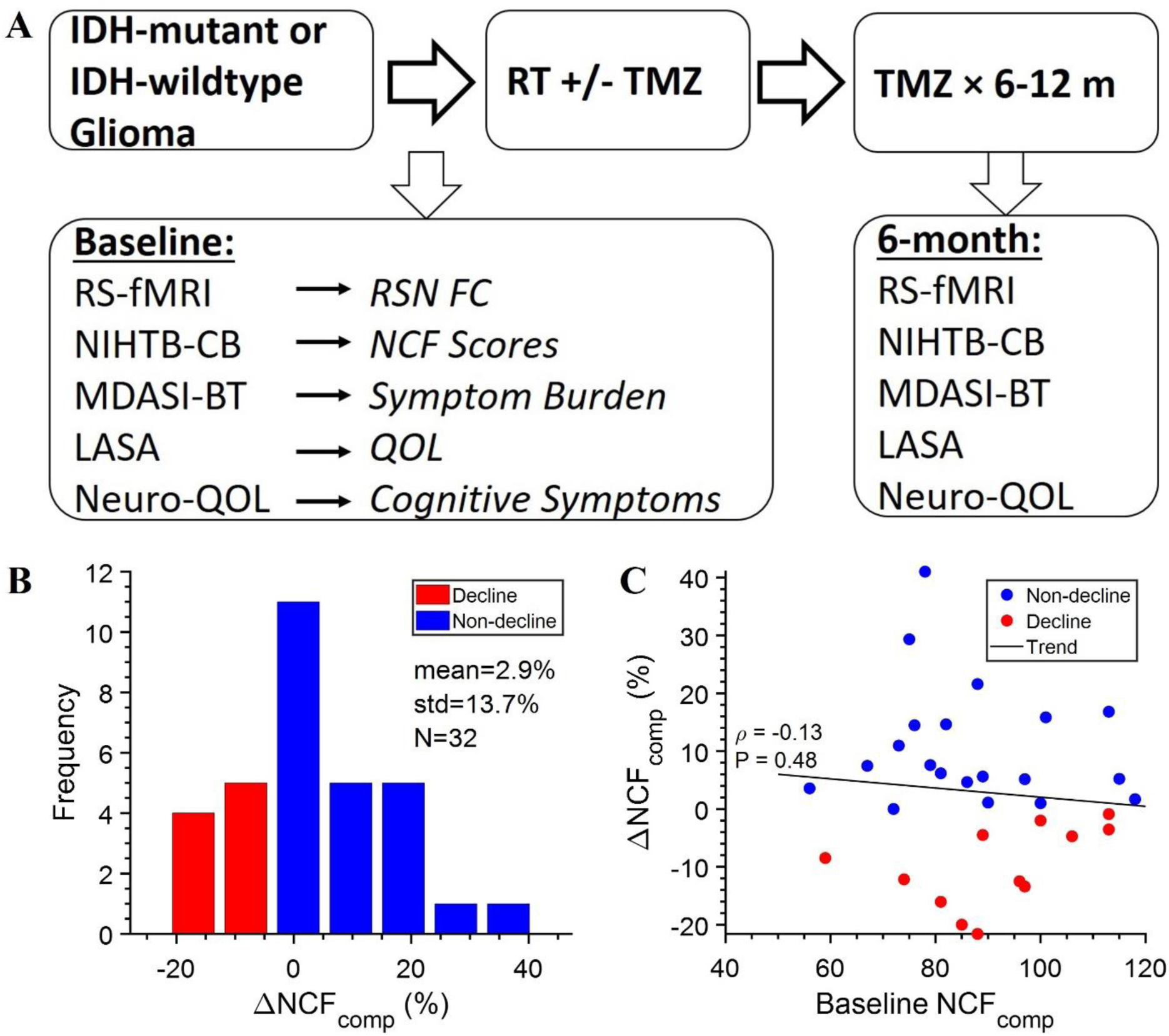
Longitudinal evaluation of neurocognitive function (NCF) and rs-fMRI changes in patients with IDH-mutant or IDH-wildtype gliomas post-RT. **(A)** Study schema. **(B)** Histogram showing the percent change in the fluid composite score (ΔNCF_Comp_) from the baseline NCF testing to 6 months post-RT. The decline cohort comprises patients with negative ΔNCF_Comp._ **(C)** Scatterplot illustrating the correlation between the baseline NCF_Comp_ and ΔNCF_comp_ at 6 months, assessed using Spearman’s ρ. ***Abbreviations***: rs-fMRI = resting-state functional MRI; RT = radiation therapy; TMZ = temozolomide; RSN = resting-state network; FC = functional connectivity; NIHTB-CB = National Institutes of Health Toolbox for the Assessment of Neurological and Behavior Function Cognitive Battery; NCF = neurocognitive function; MDASI-BT = M.D. Anderson Symptom Inventory Brain Tumor Module; LASA = linear analogue self-assessment; QOL = quality-of-life; Neuro-QOL = Neuro-Quality of Life Cognitive Function Short Form.

### Eligibility

Eligible patients must have a histological diagnosis of IDH-wildtype astrocytoma, IDH-mutant astrocytoma, or IDH-mutant oligodendroglioma, be aged 18 or older, and have a Karnofsky performance status ≥ 70. Additionally, they must be English-speaking and scheduled to receive RT (with or without chemotherapy). Given the typically poor survival of IDH-wild-type GBM patients post-chemoradiotherapy, only those with an estimated 6-month survival >80%, based on a previous validated nomogram^21^, are included. Exclusion criteria included prior cranial RT, gliomatosis, leptomeningeal disease, or medical contraindications to MRI (including those requiring anesthesia for MRI).

### Neurocognitive Testing

NCF evaluation is conducted using the National Institutes of Health Toolbox for the Assessment of Neurological and Behavior Function Cognitive Battery (NIHTB-CB), a validated, normed, and multidimensional battery of computerized NCF tests. The NIHTB-CB is designed to measure outcomes in longitudinal studies and provides age-adjusted benchmarked score normalized to the healthy population (mean of 100, SD of 15). It is freely available, relatively quick to administer on an iPad, and requires minimal training.^22–24^ We use the NIHTB-CB to examine five cognitive domains commonly affected by RT: executive function (dimension change card sort test), inhibitory control and selective attention (flanker test), episodic memory (picture sequence test), working memory (list sorting test), and processing speed (pattern comparison test). To reduce the dimensionality of the NCF performance, the five standardized test scores can also be computed into a fluid cognition composite score (NCF_Comp_), which reflects an individual’s capacity to process and integrate information, act, and solve novel and complex problems. The change of composite (ΔNCF_Comp_) is measured by the percent change from baseline to the 6-month follow-up. NCF impairment is defined when a cross-sectional standardized test score falls below 77.5 (1.5 SD below the mean).

### Patient Reported Outcomes (PROs)

The M.D. Anderson Symptom Inventory Brain Tumor Module (MDASI-BT), which includes 23 symptom and 6 interference items rated on an 11-point scale (0-10, with 10 indicating the worst symptom), was used to evaluate symptom severity and interference. The symptom severity composite score was calculated as the average of the 23 symptom items, and the symptom interference composite score as the average of the 6 interference items. The cognitive factor, one of the six sub-constructs of MDASI-BT, represented the average of the four items assessing difficulty with understanding, remembering, speaking, concentrating. The affective factor averaged eight items assessing distress, fatigue, sleep disturbance, sadness, and irritability. An increase of more than one point from the baseline indicated deterioration.^25,26^ The linear analogue self-assessment (LASA), a single-item scale ranging from 0 (worst) to 10 (best QOL), was used to evaluate self-perceived quality of life (QOL), and a decrease of one point represented deterioration.^27,28^ Following the enrollment of fifth patient, the Neuro-Quality of Life Cognitive Function Short Form Version2.0 (Neuro-QOL) was added to the PRO assessment. The eight-item questionnaire, which evaluates specific cognitive symptoms and perceived ability to complete everyday tasks, uses standardized T-scores (mean of 50, SD of 10), with lower score suggesting lower cognitive QOL. A decrease of more than 5 points from baseline on Neuro-QOL was considered deterioration.^29^

### rs-fMRI Acquisition

rs-fMRI images were obtained using dedicated 3-Tesla MRI scanners within our imaging core facility. Patients underwent imaging without sedation and were instructed to remain motionless and awake while focusing on a fixation cross. Structural images, including T1-weighted MP-RAGE and T2-weighted images, were also captured. rs-fMRI data were acquired using an echo planar imaging sequence consisting of two 6-minute runs, each with 160 frames. This sequence was sensitive to BOLD contrast and featured a repetition time (TR) of 2.2 seconds, an echo time (TE) of 30 milliseconds, and a voxel size of 3 × 3 × 3 mm^3^.

### rs-fMRI Preprocessing

rs-fMRI preprocessing was performed using seed-based correlation analysis method, as described previously.^17,30–32^ To summarize, initial processing steps were performed using the 4dfp software package developed at Washington University in St. Louis (https://4dfp.readthedocs.io/en/latest/). The BOLD data were corrected for slice timing and intensity non-uniformity consequent to interleaved acquisition. Motion censoring was conducted to exclude frames with significant movements.^33^ Spatial smoothing with a full-width at half-maximum (FWHM) of 7 mm and low-pass temporal filtering (retaining frequencies below 0.1 Hz) were also applied. Artifact was reduced using a component-based noise correction method^34^ with nuisance regressors from head motion correction, white matter, ventricles, and the global signal averaged over the whole-brain.^35^ Head motion was corrected using a rigid body transform to align each volume to the first BOLD data frame. The BOLD data then underwent a 12-parameter affine registered to the pre-radiation T1-weighted image, which was then affine-registered to the Talairach atlas.^36^ Due to the volume effects of the tumor, the registration of the T1-weighted image to the Talairach atlas was then refined with a non-linear transformation with tumor masking using Advanced Normalization Tools (ANTS) (https://www.nitrc.org/projects/ants) as previously described.^37^ This non-linear transformation was then applied to the BOLD data resampled into 3 x 3 x 3 mm³ space. Each imaging dataset underwent rigorous quality control as shown in **Supplementary Figure S1**.

### Processing of rs-fMRI to generate FC matrix

Following preprocessing, a region-of-interest (ROI) to ROI correlation matrix was computed based on 300 spherical ROIs, each with an 8-mm diameter. Each ROI was pre-assigned to one RSN as previously described.^31^ Notably, 53 subcortical ROIs have reduced correlation strength due to their distance from the coil, so they are assigned to the basal ganglia, thalamus, and cerebellum instead of specific RSNs.^31^ The BOLD timeseries for each ROI was calculated by averaging the voxel values within the ROI at each time point. A seed-based correlation map was then generated for each subject by computing the Pearson correlation between each pair of ROIs, resulting in a 300 x 300 correlation matrix (**Supplementary Figure S2**). Fisher’s r-to-z transformation was applied. ROIs within tumors, surgical cavities, subcortical regions, or exhibiting significant susceptibility-induced signal loss (>50% voxels) were excluded from the final analysis. Intra-network FC was calculated as the average correlation coefficients of ROIs within each network, represented by the on-diagonal blocks in the FC map. Inter-network FC was calculated as the average correlation coefficients of each ROI in one network to each ROI of another network, represented by the off-diagonal blocks in the FC map.

### Statistical Analysis

Patient and treatment characteristics were compared between cohorts using Fisher’s exact or Chi-squared test for categorical variables and Mann-Whitney U test for continuous variables. Correlation was assessed using Spearman’s rank correlation test (ρ). To compare the FC across ipsilateral and contralateral hemispheres, we used the paired T-test to evaluate the network-level FC difference between ipsilateral and contralateral hemispheres (i.e., for each block on the FC matrix) for each patient, where *P* > 0.05 denotes a lack of significant difference. We employed the novel connectivity regression method^38^ to select networks associated with NCF change. The whole-brain connectivity regression method utilized the Bayesian penalized multivariate regression model with the all-network FC as multivariate response and the NCF percent change as predictors. As a result, it incorporated all network FC simultaneously and avoided multiple univariate network-wise analyses, thus minimizing multiple comparison error. Permutation test was performed to identify statistically significant (*P* < 0.05) associations between network FC and NCF. The identified network FCs were then used to predict the NCF change via ordinary linear regression models.

To validate network selection and prediction performance, a split-sample approach was used for both the connectivity regression and predictive modeling analysis, with separated training set and validation set, iterated 200 times. Within each iteration, connectivity-regression analysis identified the most significant intra- or inter-network FC change associated with ΔNCF_comp_ in the training set, and linear regression predicted ΔNCF_comp_ in the validation set based on the FC changes of the selected networks, with R^2^ value used to evaluate predictive performance. Permutation test was performed 1000 times to assess the statistical significance of network selection. Initially, we empirically planned to analyze 24 evaluable cases but subsequently revised to include at least six additional patients to serve as the validation cohort, to enable the split-sample approach. Anticipating about 30% non-compliance with the 6-month NCF testing, we planned to recruit at least 43 patients to ensure 30 evaluable patients for the current analysis. By the time of the planned analysis, we had evaluable data from 32 of the 43 enrolled patients, providing a 26-patient training set and a 6-patient validation set. All statistical tests were two-sided. Statistical analyses were performed with the Statistical Package for Social Sciences version 23.0 (IBM SPSS Statistics, Chicago, IL, USA) and the R software version 4.3.1 (R Foundation for Statistical Computing, Vienna, Austria).

### Study Approval

The study was approved by the Institutional Review Board and were conducted in accordance with the Declaration of Helsinki and Good Clinical Practice guidelines. All patients provided written informed consent for participation in the study. It was registered at ClinicalTrials.gov (NCT04975139).

## RESULTS

### Patient Characteristics and Neurocognitive Function

From September 2020 to May 2023, 43 patients were enrolled in the study; 32 (74%) completed rs-fMRI and NCF tests at baseline and 6 months post-RT and were considered evaluable. Eleven patients did not complete the 6-month follow-up: 4 deaths, 5 withdraws by patient choice, one clinical decline due to disease progression, one inability to complete NCF tests. A CONSORT diagram is provided in **Supplementary Figure S3**. Baseline clinical characteristics were similar between evaluable and non-evaluable cohorts (**Supplementary Table S1**). Thirty one of the 32 evaluable patients received adjuvant RT after surgery, with the baseline rs-fMRI obtained at a median time of 1.1 months (range: 0.8 – 3.9) from surgery. One patient with IDH-mutant glioma was initially observed after surgery and developed recurrence years later, so the baseline rs-fMRI before RT was obtained 134.3 months from the initial surgery. The mean NCF_comp_ was 88.7 (SD: 16.2) at baseline and 91.1 (SD: 19.4) at 6 months, with a mean ΔNCF_comp_ of 2.9% (SD: 13.7%). Twelve patients (38%) experienced a decline in NCF_comp_, forming the “decline” cohort, while the remaining 20 patients were categorized as the “non-decline” cohort (**Figure 1B**). Clinical and treatment characteristics, including sex, education, surgical resection, tumor volume, proximity to the hippocampus, irradiated brain volume, hippocampi dose, chemotherapy, and antiepileptic medication, showed no significant differences between cohorts (Table 1). Age was the only factor with a borderline significant difference (mean 48 vs. 38; *P* = 0.11). While baseline and 6-month NCF scores across the five domains did not differ significantly different between cohorts, the decline cohort showed significantly worse changes in executive function, attention, and processing speed (**Supplementary Table S2**). ΔNCF_comp_ was not correlated with baseline NCF_comp_, suggesting that RT rather than the disease itself might be contributing more to the observed NCF decline (**Figure 1C**). The decline cohort exhibited less baseline NCF impairment than the non-decline cohort in NCF_comp_ and most individual domains (**Supplementary Figure S4A**). However, this trend reversed at 6 months, with the decline cohort showing greater NCF impairment (**Supplementary Figure S4B**). Excluding the 5 patients (all with IDH-wildtype gliomas) who showed radiological progression at 6 months, 10 progression-free patients (37%) still exhibited NCF_comp_ decline, including 7 out of 21 patients with IDH-mutant gliomas and 3 out of 6 with IDH-wild-type gliomas. Changes in NCF were not different between IDH-wildtype and IDH-mutant patients, both for the entire cohort and for the subgroup without radiological progression (**Supplementary Figure S5**).

**Table 1:**
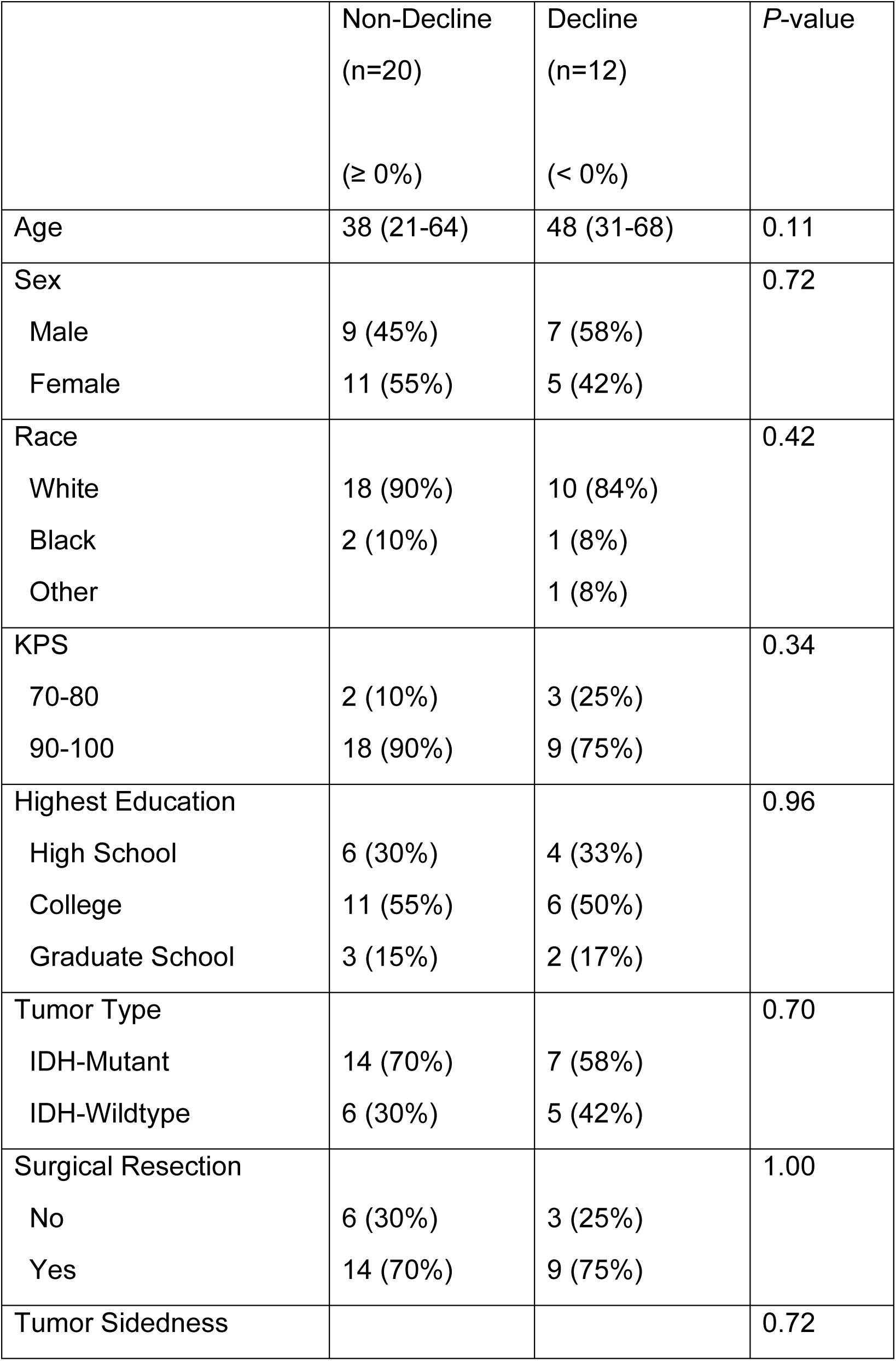
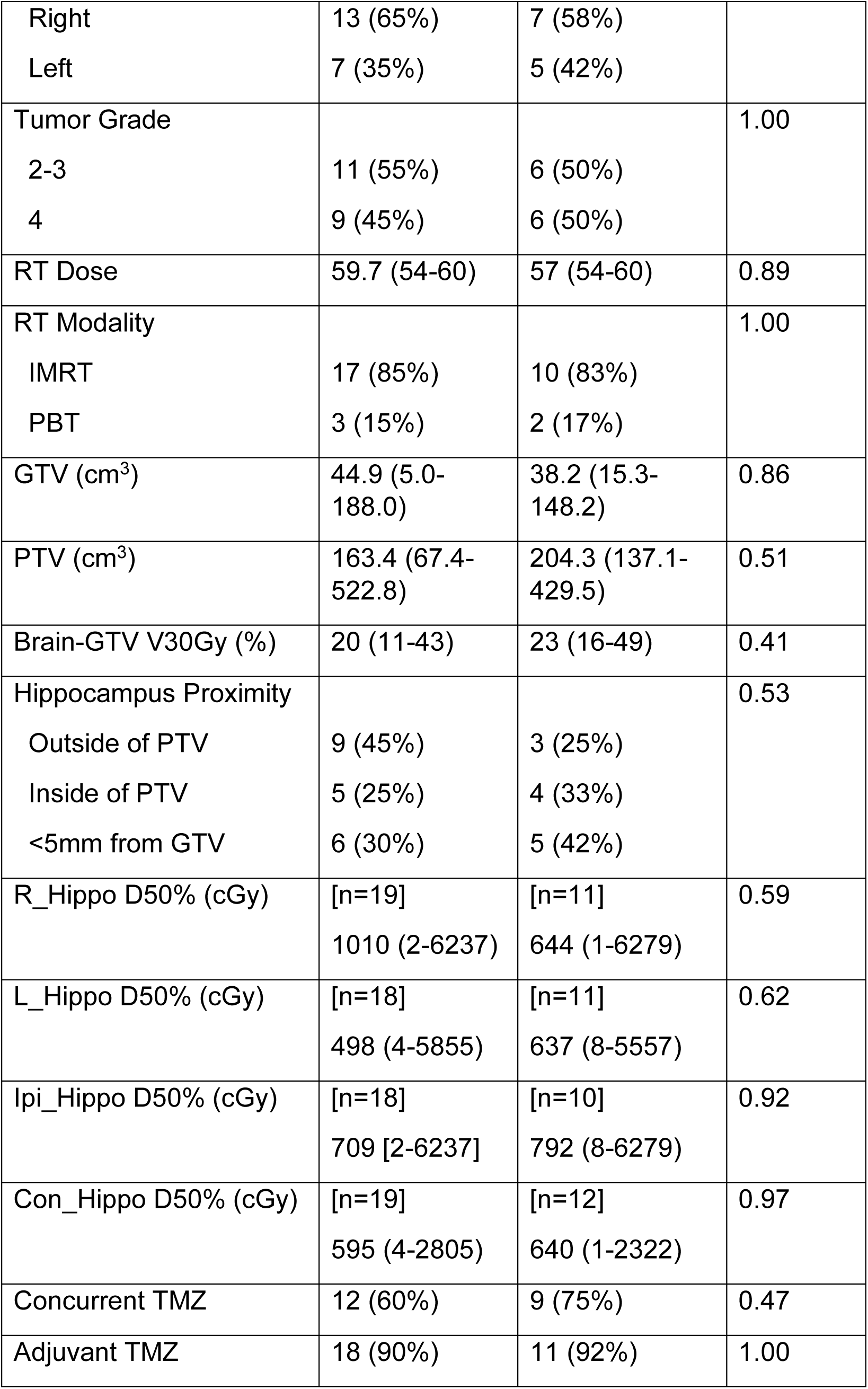
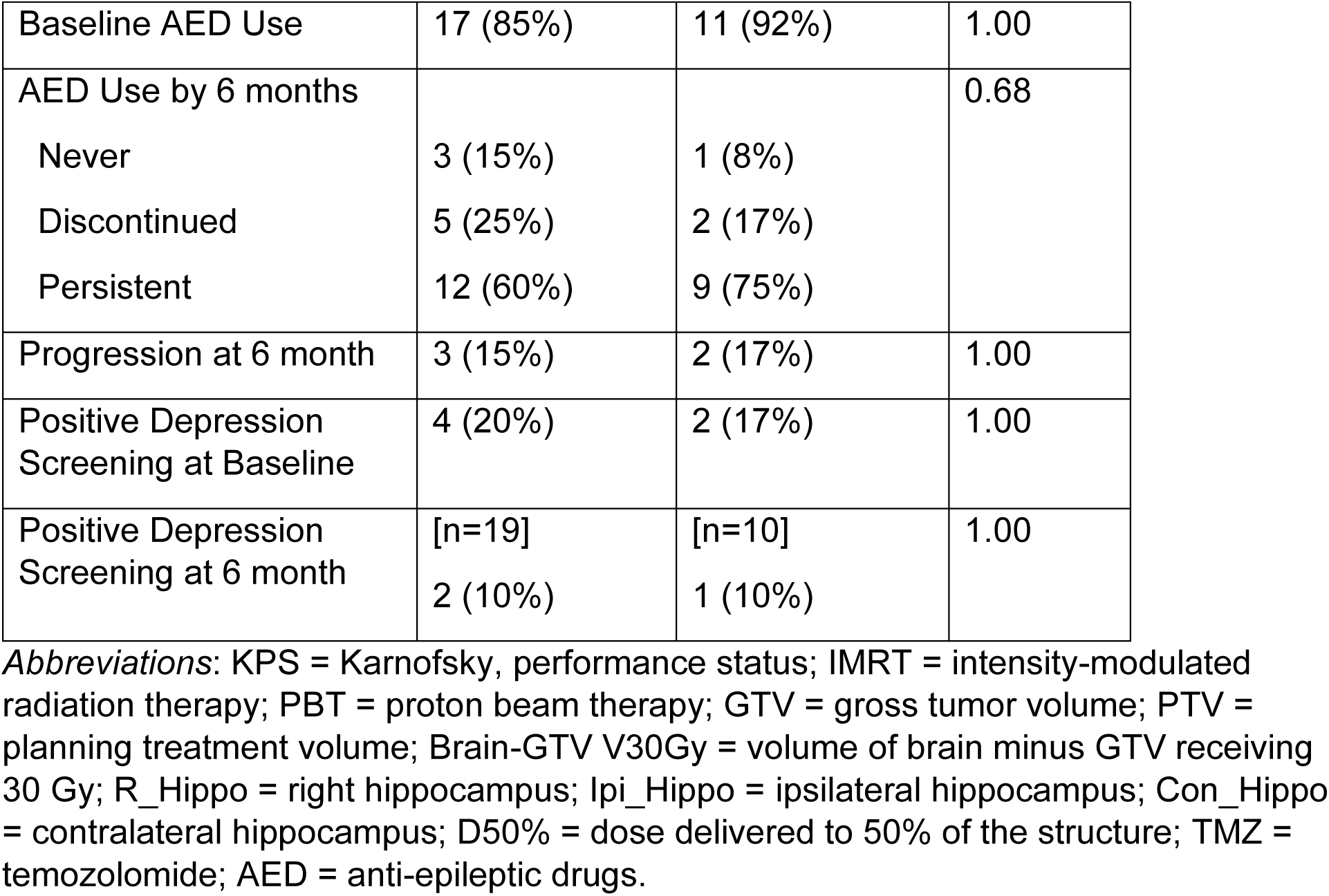
Patient characteristics of patients without NCF decline versus those with decline.

### Correlation between PRO and NCF changes

Twenty-nine patients (91%) completed the MDASI-BT and LASA at baseline and 6 months. Of these, 28% reported deterioration in symptom severity, 13% in symptom interference, 22% in cognitive factor, 25% in affective factor, and 28% in overall QOL. Twenty-seven patients (84%) completed the Neuro-QOL, with 13% reporting deterioration in cognitive symptoms. There were no significant differences in baseline, 6-month, and changes in PROs between the decline and non-decline cohorts (**Supplementary Table S3, Supplementary Figure S6A-E**). Additionally, there was no significant correlation between NCF changes versus changes in PROs (**Supplementary Figure S6F-J**), although a weak correlation was observed between NCF_comp_ and MDASI-BT cognitive factor (ρ = −0.24, *P* = 0.20, **Supplementary Figure S6H**).

### rs-fMRI Changes after RT

Composite FC matrices averaged over the 32 evaluable patients at baseline and 6 months are presented in **Figure 2A-B**. The corresponding FC matrices from the ipsilateral (same side as the tumor) and contralateral ROIs are shown in **Figure 2C-D** and **Figure 2E-F**, respectively. Despite significant tumor involvement, surgical manipulation, and RT on the ipsilateral side, intra-hemispheric FC appears overall similar between the ipsilateral and contralateral brain at baseline (**Figure 2G**) and 6 months (**Figure 2H**). Analyzing each individual separately, 29 of 32 patients (91%) had non-significantly different intra-hemispheric FC between two hemispheres at baseline, and 21 of 32 patients (66%) had non-significant difference at 6 months (**Supplementary Table S4**).

**Fig. 2.**
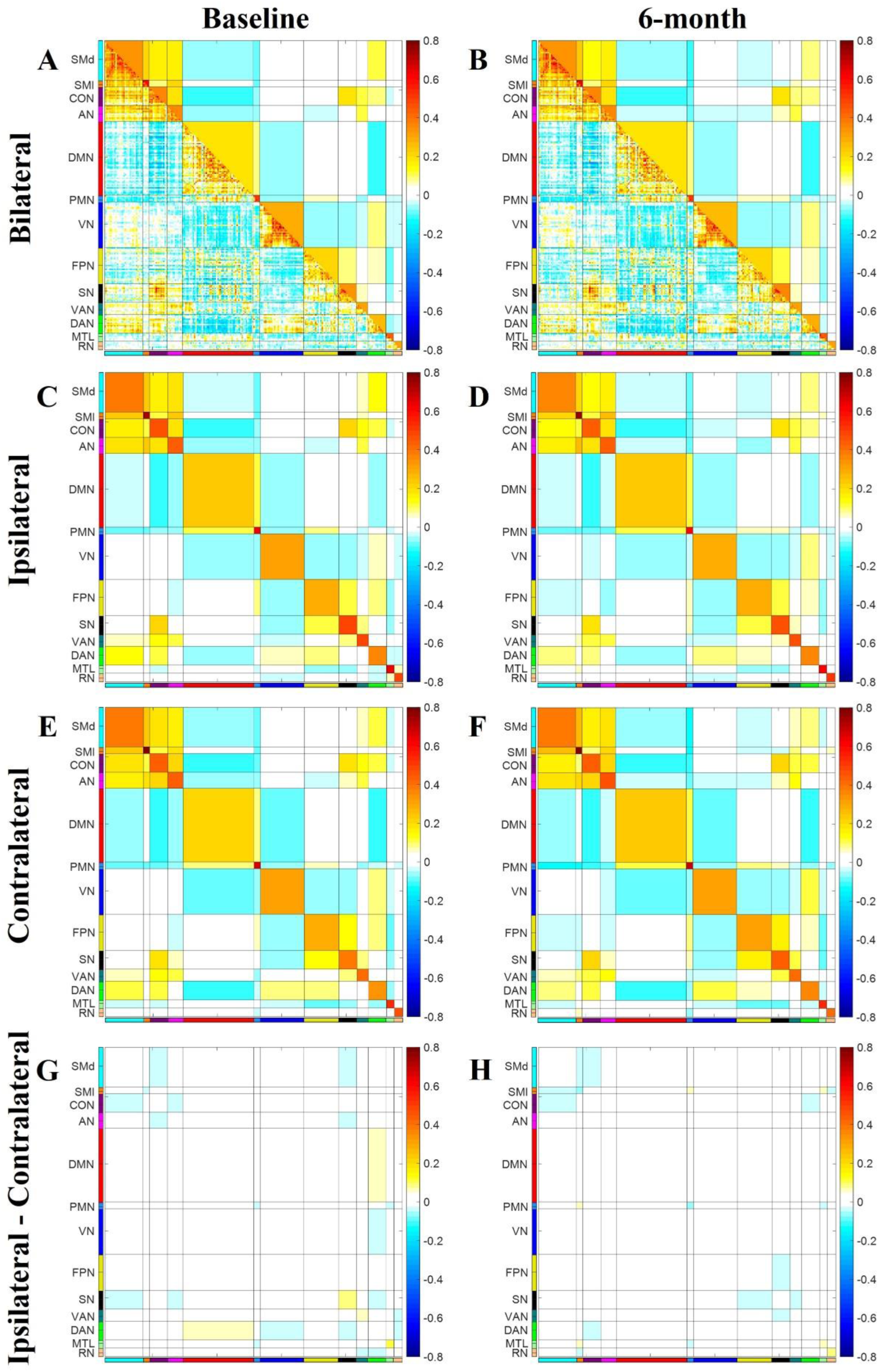
Composite functional connectivity (FC) matrices of rs-fMRIs from all patients (n = 32) before and post-RT. **(A)** Composite FC matrix (the combination of all the correlation matrices from all 32 patients) at baseline. **(B)** Composite matrix at 6 months post-RT. **(C)** Difference composite matrix of 6 months minus baseline, showing the change from baseline to 6 months post-RT. **(D-F)** Composite matrix for the ipsilateral hemisphere (same side as the tumor) at baseline, 6 months, and 6 months minus baseline, by averaging only correlation coefficients of the ROIs from the ipsilateral hemisphere. **(G-I)** Composite matrix for the contralateral hemisphere (opposite side of the tumor) at baseline, 6 months, and 6-months minus baseline, by averaging only correlation coefficients of the ROIs from the contralateral hemisphere. The bottom triangle of the correlation matrix shows the correlation coefficients between regions-of-interest (ROIs). The blocks in the upper triangle of the matrix represent the average of correlations coefficients between ROIs. The on-diagonal blocks represent the average of correlation coefficients of ROIs within each network (intra-network FC). The off-diagonal blocks represent the average correlation coefficients of each ROI in one network to each ROI of another network (inter-network FC). ***Abbreviastions***: FC = functional connectivity; SMd = dorsal somatomotor (network); SMl = lateral somatomotor (network); CON = cinguloopercular network; AN = auditory network; DMN = default mode network; PMN = parietal memory network; VN = visual network; FPN = frontoparietal network; SN = salience network; VAN = ventral attention network; DAN = dorsal attention network (DAN); MTL = medial temporal lobe network; RN = reward network.

### Correlation between rs-fMRI changes and NCF changes

As seen in **Figure 3A-F**, changes of network FC appear to differ between the non-decline and decline cohorts. Connectivity-regression analysis demonstrated moderately strong predictive performance for ΔNCF_comp_ using changes in network FC (R^2^ = 0.301, SD = 0.249). The most predictive measures were change in intra-network FC of the Parietal Memory Network (PMN-PMN) and inter-network FC between the PMN and the Visual Network (PMN-VN), which were also highly significant on permutation tests (P = 0.001 and P = 0.002, respectively; **Table 2**). Across the entire cohort (combining both training and validation sets), changes in PMN-PMN and PMN-VN were strongly correlated with ΔNCF_comp_ (ρ = 0.50 and ρ = −0.43, respectively; **Figure 4**). These findings appeared visually distinct on the composite FC results (note the contrast between the decline vs non-decline cohort; **Figure 3**). Sensitivity analysis using only contralateral ROIs yielded similar overall predictive performance and confirmed the same leading predictive measures (**Table 2**). FC matrices derived from the contralateral ROIs are displayed in **Supplementary Figure S7**.

**Fig. 3.**
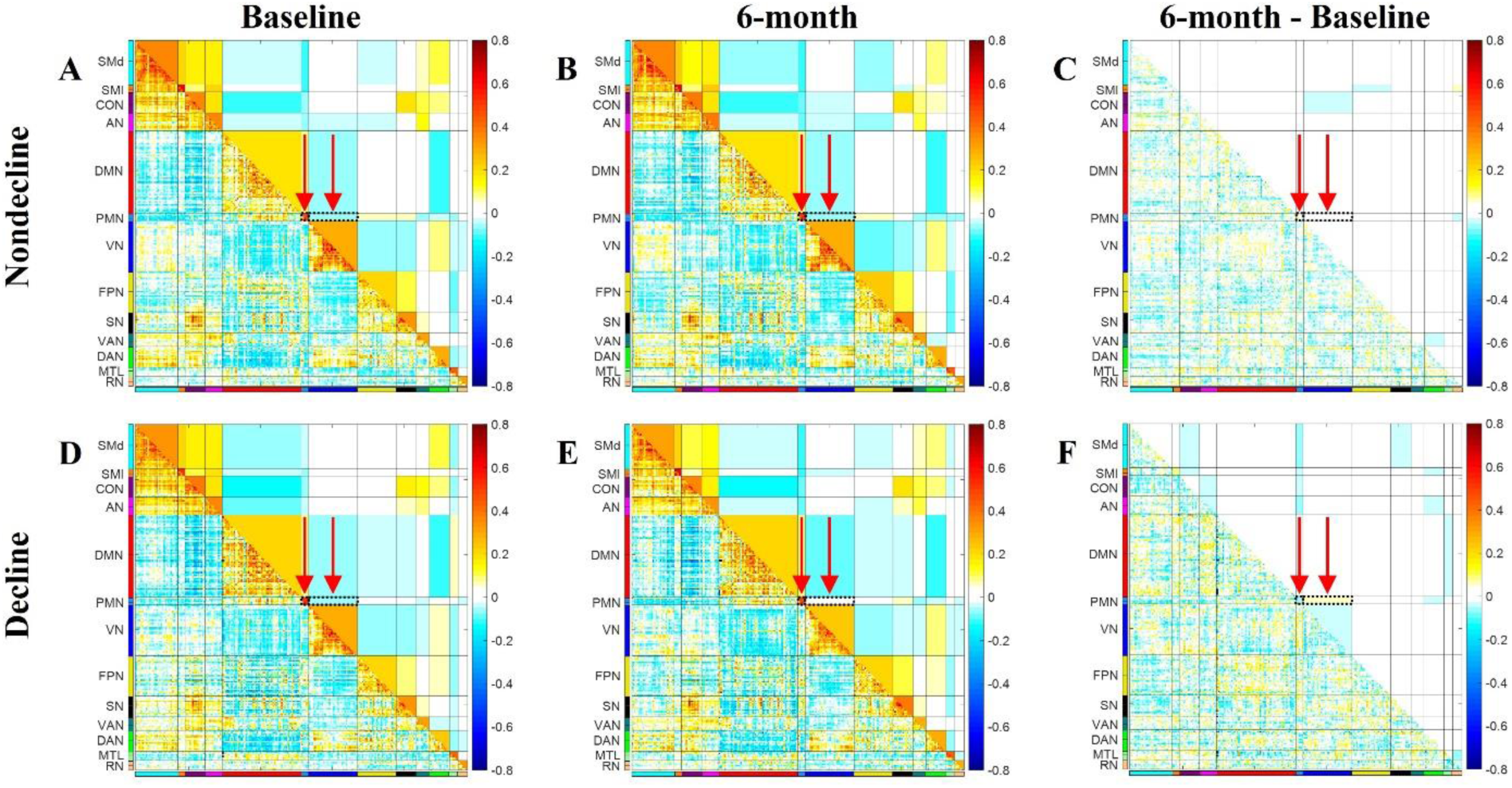
Functional connecitvity (FC) matrices for patients with and without NCF decline. **(A-C)** Composite FC matrices for the non-decline cohort (n = 20) at baseline, 6 months, and 6 months minus baseline. **(D-F)** Composite FC matrices for the decline cohort (n = 12) at baseline, 6 months, and 6 months minus baseline. Arrows indicate blocks with the strongest predictive performance for the change of NCF from the connectivity-regression analysis.

**Fig. 4.**
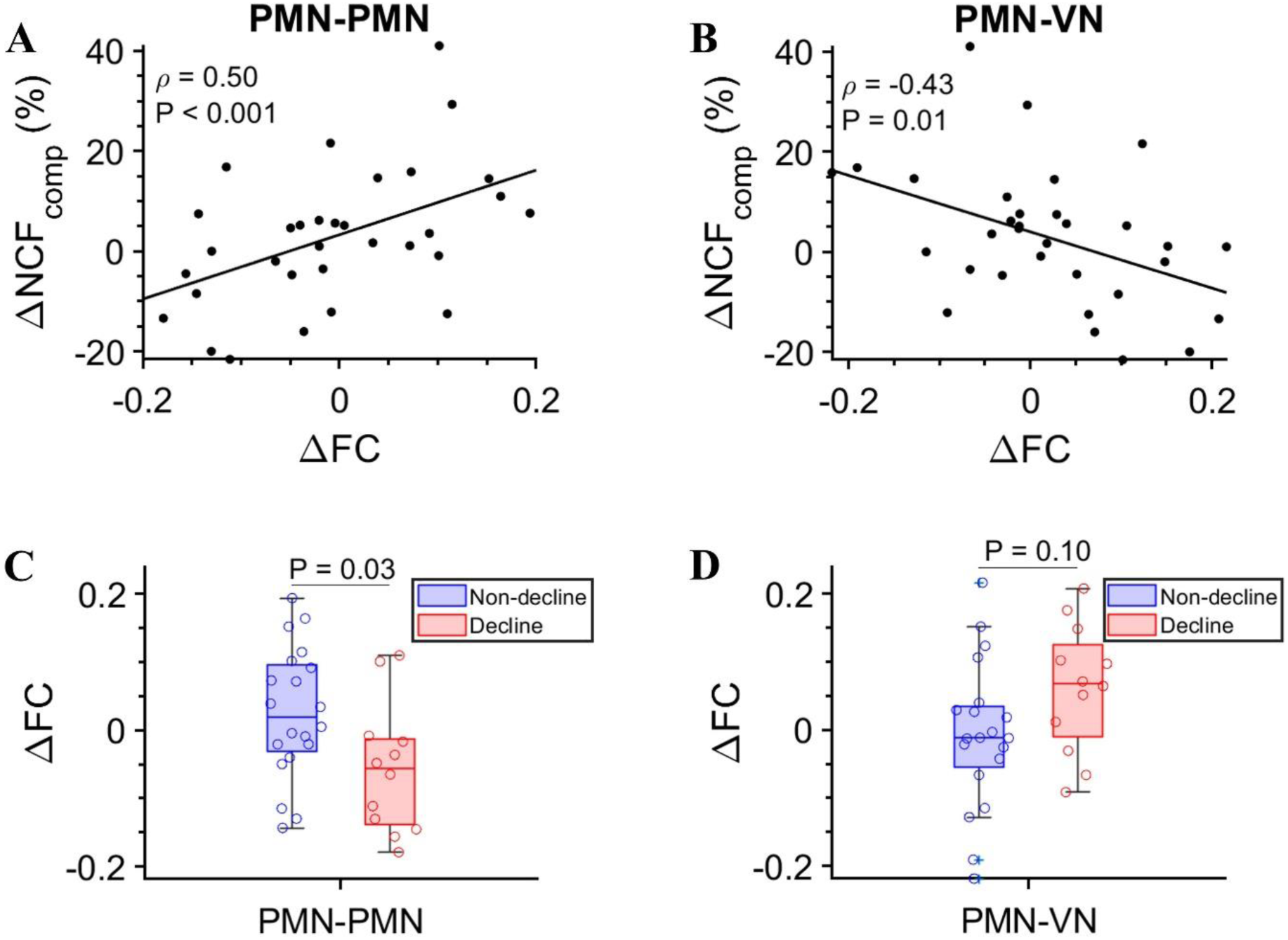
Correlation of FC changes with NCF changes in glioma patients post-RT. **(A)** Scatterplot showing the correlation between the intra-network FC change of parietal memory network (PMN-PMN) from baseline to 6 months with the percent change in NCF composite score (ΔNCF_Comp_). **(B)** Scatterplot illustrating the correlation between the inter-network FC change between PMN and visual network (PMN-VN) from baseline to 6 months with the percent change in NCF composite score (ΔNCF_Comp_). **(C)** Boxplot comparing the FC change of PMN-PMN between the decline and non-decline cohorts. **(D)** Boxplot comparing the FC change of PMN-VN between the decline and non-decline cohorts. Correlation was evaluated using as Spearman’s ρ in A and B; p-values were determined by Mann-Whitney U test in C and D.

**Table 2:** Connectivity regression analysis using 200 training sets (n = 26 patients) and 200 validation sets (n = 6 patients)

| Analysis Type | Variable Used | R<sup>2</sup> from 200 validation sets [mean, (SD)] | The top variables selected by >50% among 200 training sets | P-value* |
| --- | --- | --- | --- | --- |
| Primary analysis with bilateral rs-fMRI maps of all RSNs | Intra-network and inter-network FC changes of all 13 RSNs (n = 91) | 0.301 (0.249) | PMN-PMN (100%) | 0.001 |
|  |  |  | PMN-VN (99%) | 0.002 |
|  |  |  | AN-AN (81.5%) | 0.002 |
|  |  |  | DAN-SMI (76%) | 0.010 |
|  |  |  | PMN-AN (51.5%) | 0.035 |
| Sensitivity Analysis with only contra-lateral rs-fMRI maps of all RSNs | Intra-network and inter-network FC changes of all 13 RSNs (n = 91) | 0.259 (0.235) | PMN-VN (100%) | <0.001 |
|  |  |  | PMN-PMN (99.5%) | 0.001 |
|  |  |  | DMN-VAN (59%) | 0.014 |
|  |  |  | CON-VN (51.5%) | 0.002 |

\*P-values were determined using permutation test, iterated 1000 times.
*Abbreviations:* rs-fMRI = resting state functional MRI; RSN = resting-state networks; FC = functional connectivity; intra-network FC of parietal memory network = PMN-PMN; inter-network FC between PMN and visual network = PMN-VN; AN = auditory network; SMI = lateral somatomotor (network); DAN = dorsal attention network (DAN); DMN = default mode network; VAN = ventral attention network; CON = cinguloopercular network.

## DISCUSSION

We successfully conducted a longitudinal, prospective study of rs-fMRI, NCF, and PROs assessments in adult patients with either IDH-wildtype or IDH-mutant gliomas before and 6 months after partial-brain irradiation. We observed approximately 38% patients developed NCF decline post-RT. There was no significant difference in NCF changes between IDH-wildtype versus IDH-mutant patients, nor was there a significant correlation between NCF changes and subjective QOL changes. Despite the significant impact of tumor, surgery, and RT to one hemisphere, FC of the contralateral side was comparable to the ipsilateral side for majority of patients, both before and after RT. Overall, FC changes in rs-fMRI accounted for a moderate amount of variance in NCF changes (R^2^ = 0.301), with PMN-PMN and PMN-VN FC changes emerging as the most robust predictors of cognitive decline.

To our knowledge, few prospective studies have simultaneously conducted NCF testing and rs-fMRI in patients with diffuse gliomas before and after chemoradiotherapy. Although several important prospective studies have performed NCF testing in this context^11,39,40^, none included concurrent rs-fMRI. These studies generally found that only a subset of glioma patients develop statistically significant NCF decline after RT, with the majority of progression-free patients showing stable or improved NCF post-RT, which aligns with our observation. Koche et al. conducted a cross-sectional study involving rs-fMRI and NCF testing in 80 glioma patients, primarily referred for advanced imaging to monitor residual tumors post-RT or suspected recurrence post-RT. The median interval between therapy initiation and imaging was 13 months (range: 1–114), with 73% having completed chemoradiotherapy. Their analysis, which was limited to the Default Mode Network (DMN), found decreased FC in selected DMN nodes that were associated with decreased NCF compared to healthy controls.^41^ However, that study was not longitudinal and included a heterogenous patient population at various stages of therapy or recovery. A prospective study conducted rs-fMRI and NCF testing for 39 patients with nasopharyngeal tumor before and 3 months after RT, which typically would irradiate the inferior temporal lobes. Aside from a different patient population, the study also used a less sensitive NCF instrument (the Montreal Cognitive Assessment) and only analyzed three RSNs (DMN, Executive Control Network [ECN], and Salience Network [SN]). The study observed decreased intra-network as well as inter-network FC 3 months after RT, but no significant associations between FC changes and NCF decline.^42^

Although IDH-wildtype gliomas are more resistant to chemoradiotherapy than IDH-mutant gliomas, it remains unclear whether the impact of chemoradiotherapy on NCF changes differs between these different histological tumor types, especially in the absence of tumor progression, a known factor in NCF decline.^43,44^ Our data indicate similar NCF changes between IDH-mutant gliomas and (favorable-prognostic) IDH-wildtype gliomas at least 6 months after chemoradiotherapy. In RTOG-0825, a landmark randomized study primarily involving IDH-wildtype GBM patients (approximately 95%), longitudinal NCF testing was conducted using a different battery of tests for patients without clinical or radiological signs of progression.^11^ That study reported a 36% decline in NCF composite scores at 6 months after RT and TMZ, comparable to the 33% observed in our cohort of 21 progression-free IDH-mutant glioma patients. NRG-BN005, a recently completed randomized phase II study comparing proton-based RT vs photon-based RT in IDH-mutant gliomas, also used the same NCF test battery as RTOG-0825 and should provide further insights into the incidence of radiation-related NCF decline in this patient group (NCT03180502).

Our finding that objective NCF testing does not strongly correlate with subjective PROs aligns with existing literature. Patient-reported PROs assessments provide complementary information to objective assessment and can provide a more comprehensive view on the impact of NCF changes.^45^ However, subjective cognitive concerns can be influenced by mood, a lack of insight into one’s own deficits, and variability in self-perception, leading to typically weak correlations between objective NCF performance and self-reported cognitive function among most studies.^45,46^ Consequently, objective NCF testing is typically considered the gold standard for evaluating the impact of therapies on NCF.

To our surprise, we observed that FC of the ipsilateral side of the brain in glioma patients, as derived from rs-fMRI, is similar to the contralateral side before and after RT, despite considerable impact from the tumor, surgery, and RT. This phenomenon has also been reported in other studies. Mallela et al. compared rs-fMRI in 24 glioma patients with 12 healthy controls, focusing on FC between primary and supplementary motor areas. They identified a similar reduction in FC across both hemispheres, unaffected by proximity to the tumor.^47^ Cho et al. examined the left and right language RSNs in 29 glioma patients with left-hemispheric brain tumors and found comparable reductions in FC across both ipsilateral and contralateral hemispheres compared to healthy controls.^48^ In another rs-fMRI imaging study assessing global anomalies of FC patterns in 15 GBM patients compared to healthy controls, Nenning et al. also noted that network anomalies were highly symmetric across both hemispheres, regardless whether they were ipsilateral or contralateral to the tumor.^49^ Analyzing 189 patients with newly diagnosed GBM and 189 age-matched healthy controls, Park et al. observed whole-brain spectral slope flattening of the BOLD signal fluctuations, that was comparable between ipsilateral and contralateral hemispheres. This spectral flattening suggested widespread neuropathological disruption across the entire brain.^50^ Hadjiabadi et al. conducted a preclinical rs-fMRI study comparing tumor-bearing to healthy murine brains and observed global attenuation of FC that extended to the contralateral hemisphere. Histological analysis indicated that these brain-wide alterations in rs-fMRI signals were due to tumor-related neurovascular remodeling.^51^ In another preclinical study, Seitzman et al used wide field optical imaging (WFOI) to simultaneously measure neural and hemodynamic signaling before and after whole-brain RT of tumor-free mice. They also observed widespread RSN FC changes associated with concomitant reduced neuronal activity but not hemodynamic activity.^52^ Collectively, these results suggest that gliomas may induce global changes in RSN connections, indicating a brain-wide rather than focal disruption of the underlying neural process. Our similar findings post-RT may also suggest that the effect of partial-brain irradiation on neuronal activity are also widespread and not merely regional.

Our analysis demonstrates that changes in rs-fMRI after chemoradiotherapy significantly predicted NCF changes, suggesting that rs-fMRI may be a promising imaging biomarker of NCF decline. We identified specific regions within rs-fMRI, such as PMN-PMN and PMN-VN, as particularly sensitive to detect NCF changes post-RT. The PMN, a RSN identified through meta-analysis using task-based fMRI and rs-fMRI, is anatomically adjacent to the DMN and VN.^53^ It plays a unique role in learning and memory, showing increased activation with stimulus repetition and facilitating hippocampus-independent memory processing of new information.^54,55^ Strong FC between PMN and VN was detected in experiments in which participants underwent rs-fMRI while watching movies, indicating that communication between these networks is crucial for memory processing.^56^ We hypothesize that patients with radiation-induced cognitive decline may exhibit decreased neural connections within the PMN and with the VN, as reflected in rs-fMRI.

Major limitations of our study include a modest sample size and the potential for multiple-testing error due to the large number of variables derived from rs-fMRI (91 network-level FC changes). To mitigate these issues, we used the novel connectivity-regression method that incorporates all network FC simultaneously to avoid multiple univariate network-wise analyses, thus minimizing multiple comparison error. We also employed a split-sample approach with 200 iterations of training set and validation set, along with one thousand runs of permutation tests. Despite these complex statistical models, the potential for false discovery remains owing to an extensive data space. Consequently, external validation with a larger patient cohort is necessary in the future. Our study suggests strong associations between specific RSN FC changes on rs-fMRI and NCF changes, but these findings do not imply causation. It is possible that these regions are simply more sensitive to the global disorganization of RSNs following radiation injury. Further research is required to determine whether specific injuries to the neural connections in PMN-PMN and PMN-VN directly contribute to radiation-induced cognitive decline. Future research should also explore the potential clinical applications of rs-fMRI as an imaging biomarker, including early detection of radiation-related neurocognitive decline and providing a platform to evaluate the efficacy of novel strategies aimed at reducing this decline.

## Data Availability

De-identified clinical data from this prospective clinical trial will be available upon reasonable request to the authors.

## Acknowledgments

We thank Karen Miller, Konstantina Stavroulaki, David Schwab, Anna Antiporda, Charlotte Phillips, and Stephanie Myles in the Department of Radiation Oncology for clinical trial management. We thank the Alvin J. Siteman Cancer Center at Washington University School of Medicine and Barnes-Jewish Hospital in St. Louis, MO, for the use of the Shared Resources including Clinical Trials Core and the Center for Clinical Imaging Research facility. The Siteman Cancer Center is supported in part by an NCI Cancer Center Support Grant #P30 CA091842. We thank the Department of Radiation Oncology for shared resources.

## Author contributions

Zhihua Liu and Tim Mitchell performed research, analyzed data, and wrote the manuscript. Chongliang Luo and Ki Yun Park performed research, analyzed data, and reviewed the manuscript. Joshua Shimony, Robert Fucetola, Abraham Snyder, and Tong Zhu designed studies, analyzed data, and reviewed the manuscript. Eric Leuthardt and Stephanie Perkins performed research, analyzed data, and reviewed the manuscript. Jiayi Huang designed studies, performed research, analyzed data, supervised the study, and wrote the manuscript.

## Data availability

De-identified clinical data from this prospective clinical trial will be available upon request.

