## Supplemental Figures and Tables for "Neurocognitive and resting-state functional MRI changes in patients with diffuse gliomas after chemoradiotherapy"

### Supplementary Figures and Tables

#### Supplementary Figure S1: An example quality control report for rs-fMRI preprocessing.

**(A)** The movement analysis for the two BOLD runs. On the x-axis, frame numbers are plotted, with frames 1-160 corresponding to the first BOLD run and frames 161-320 to the second. The y-axis represents the motion parameters, including three translational and three rotational movements. **(B-C)** Axial cross-sectional images through the middle of the brain (left) and through the middle of the tumor (right), with the underlying image being the standard atlas and the top green borders representing the patient's gray and white matter registered in the standard space. Dice coefficient or Dice similarity index was used to evaluate similarity between segmented patient's gray matter versus that of the standard atlas, serving as a metric for image registration accuracy. **(D)** The spatial standard deviation of successive difference images (DVARS) over time for the two BOLD runs (with red dots marking frames censored by the format file). **(E)** Histogram displaying the distribution of DVARS values, with values below the critical threshold colored green and those above in red. **(F)** Functional connectivity (FC) matrix based on the correlation coefficient of 36 selected ROIs. **(G)** Seed maps for four selected ROIs: ROI_1 (posterior cingulate cortex), ROI_3 (left lateral parietal), ROI_30 (left motor cortex), and ROI_31 (right motor cortex). These ROIs were selected due to their well-established FC pattern in the brain networks, particularly in relation to cognitive and motor functions. The seed maps illustrate the FC between each selected ROI and other brain regions. In these maps, regions with positive correlations are functionally connected with the seed ROI, indicating synchronous activity, while regions with negative correlations may indicate anti-correlated activity.

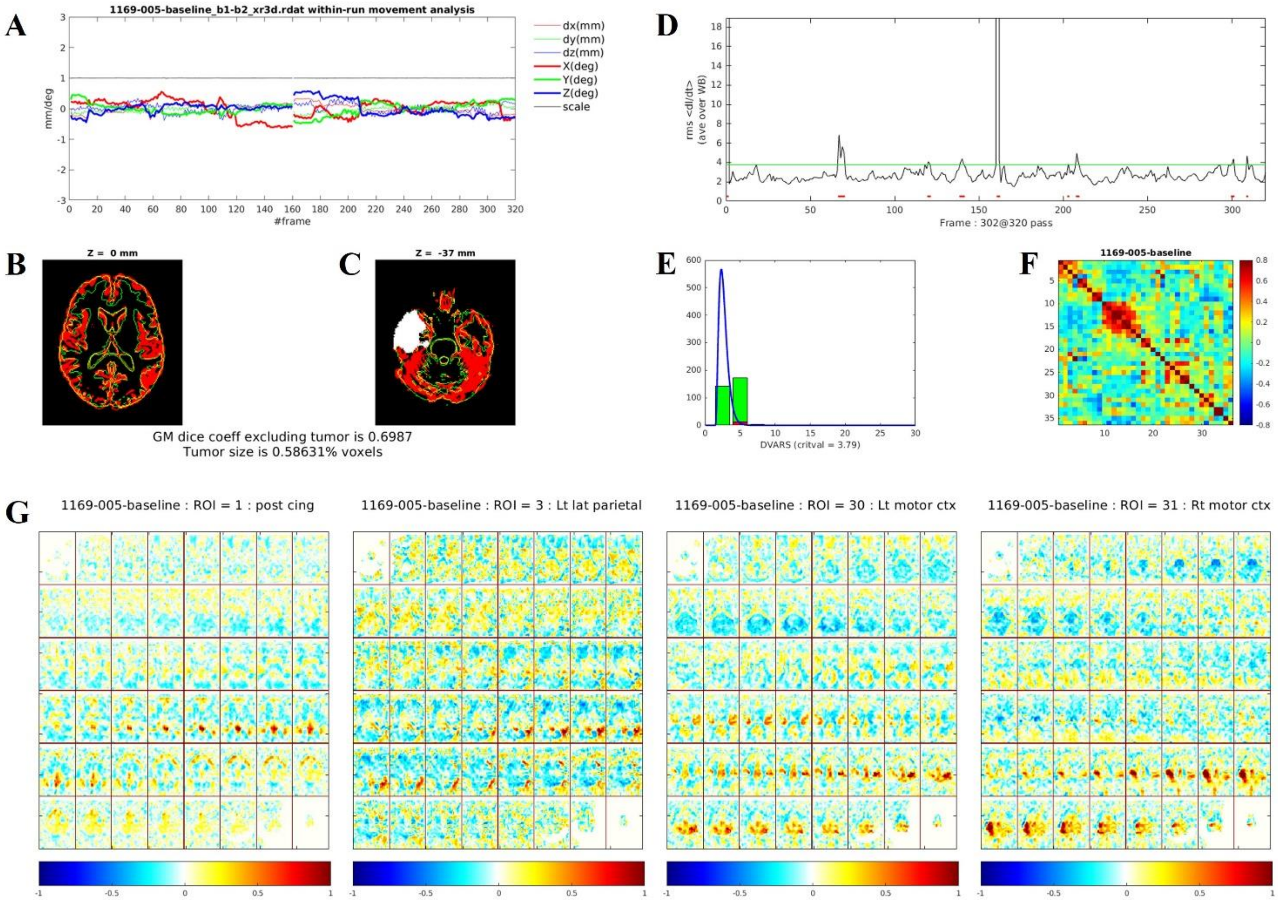

**Supplementary Figure S2: Functional connectivity (FC) matrix of rs-fMRI.** FC matrix map is generated by computing the Pearson correlation between each pair of 300 spherical regions-of-interest (ROIs), resulting in a 300 x 300 correlation matrix. These ROIs were pre-assigned to 14 resting-state networks (RSNs) and 3 subcortical regions (basal ganglia, thalamus, and cerebellum). The bottom triangle of the correlation matrix shows the raw correlation coefficients between ROIs. The blocks in the upper triangle of the matrix represent the average of correlations coefficients between ROIs. The on-diagonal blocks represent the average of correlation coefficients of ROIs within each network (intra-network FC). The off-diagonal blocks represent the average correlation coefficients of each ROI in one network to each ROI of another network (inter-network FC). Solid black lines in the bottom triangle represent ROIs that were excluded due to tumor mask or susceptibility-induced signal loss.

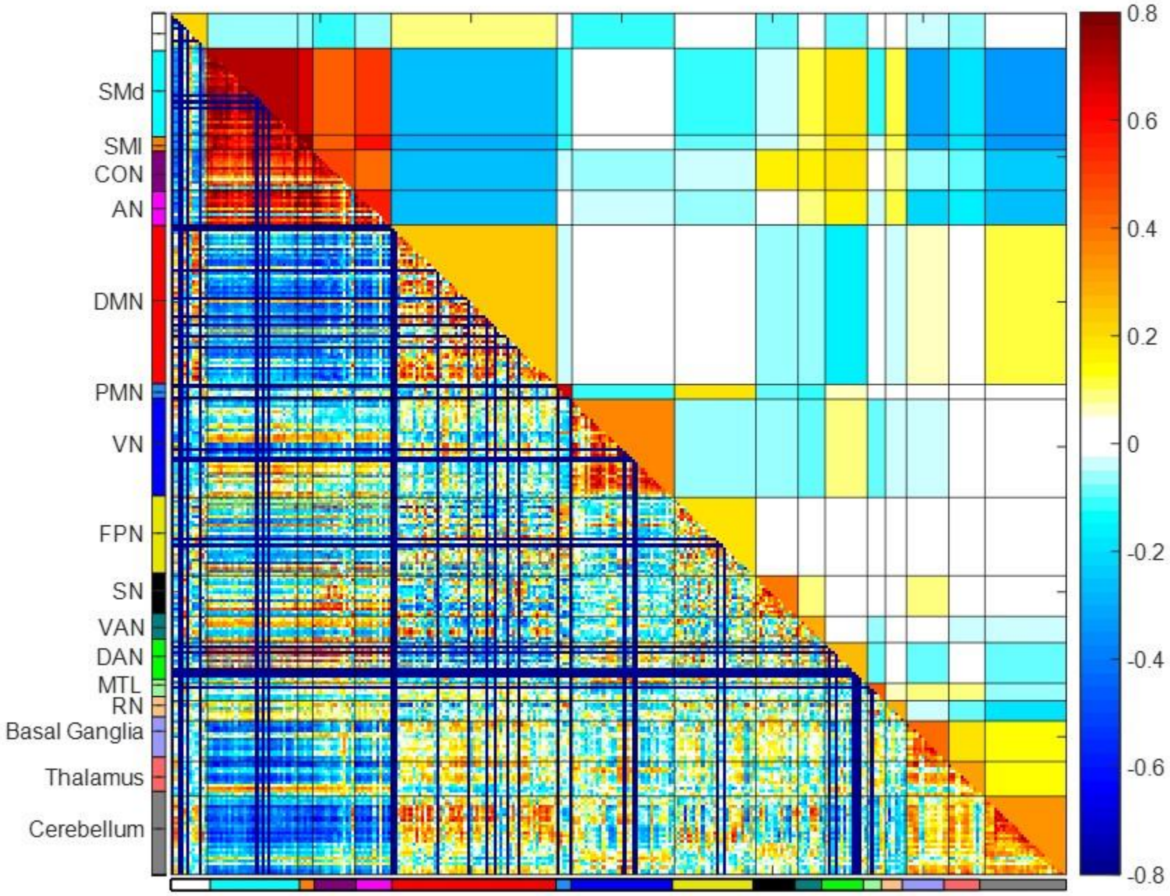

**Supplementary Figure S3: CONSORT flow diagram of the study.**

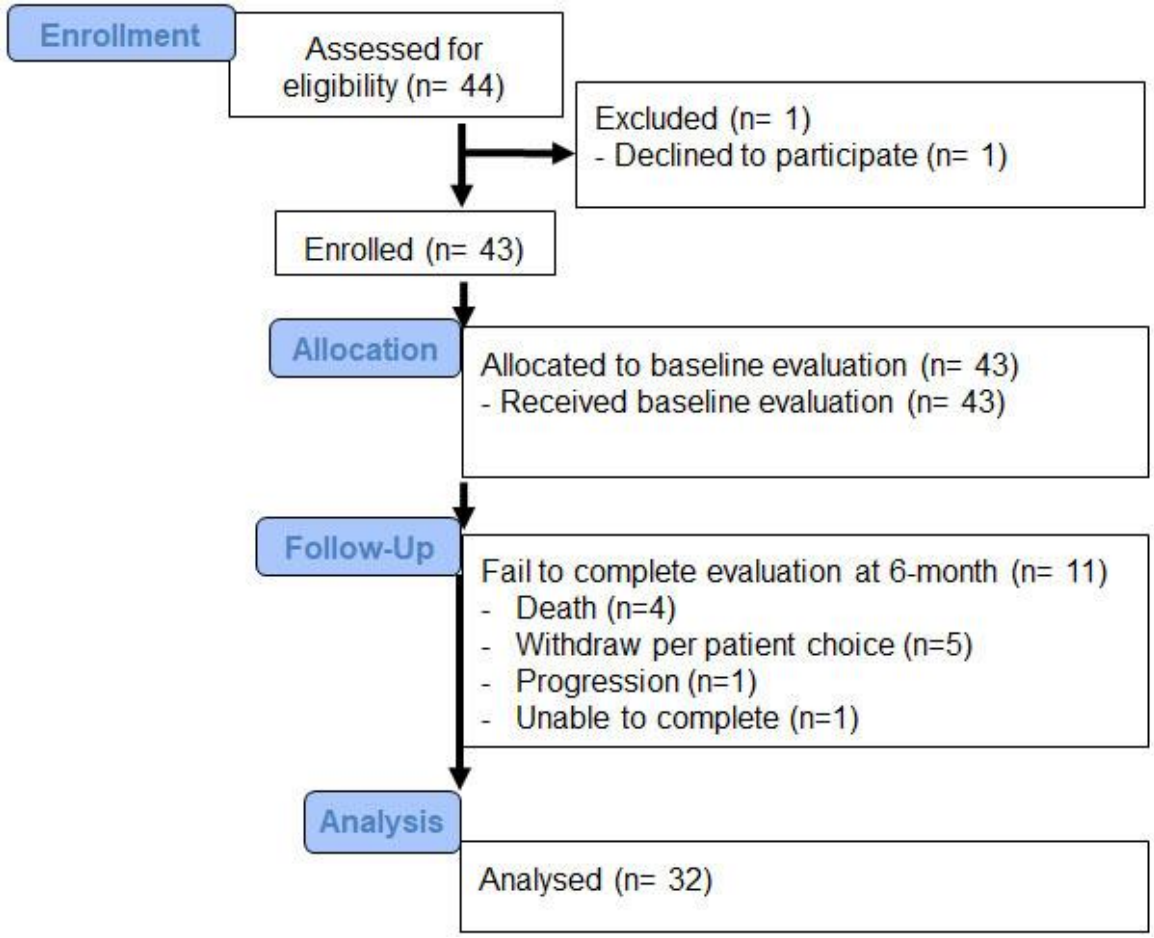

**Supplementary Figure S4: Neurocognitive function (NCF) impairment across neurocognitive tests. (A)** Bar plot showing the NCF impairment rates at baseline across the fluid composite score and each of the five specific cognitive tests. NCF impairment is defined as a score below 77.5 (1.5 SD below the mean). **(B)** Bar plot showing the NCF impairment rates at 6 months across the fluid composite score and each of the five specific cognitive tests. *Abbreviations*: $NCF_{composite}$ = fluid cognition composite score computed from the five cognitive tests; EF = executive function (dimension change card sort test); ICSA = inhibitory control and selective attention (flanker test); EM = episodic memory (picture sequence test); WM = working memory (list sorting test); PS = processing speed (pattern comparison test).

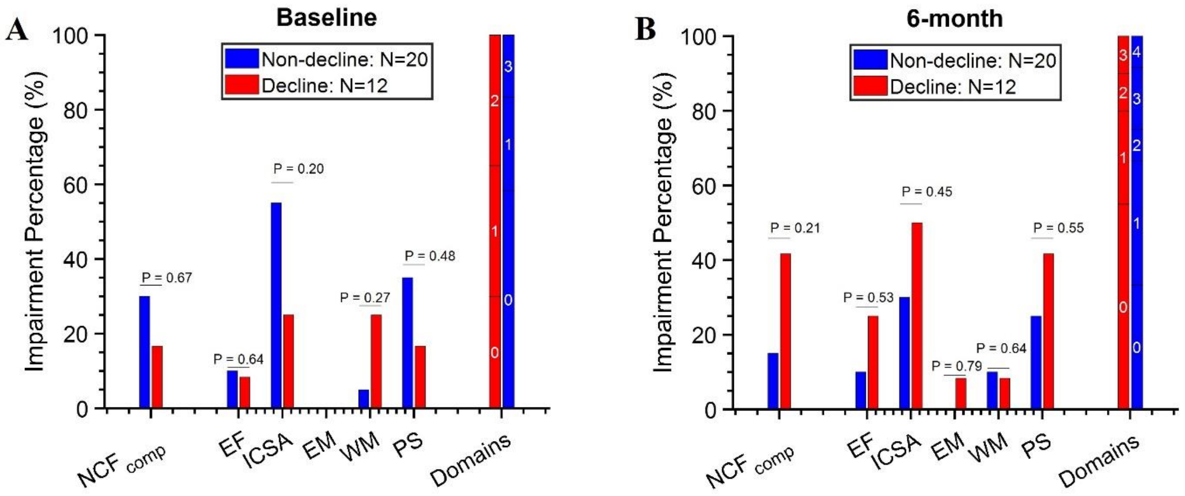

**Supplementary Figure S5: Comparison of NCF changes between IDH-mutant and IDH-wildtype glioma patients.**
**(A)** all patients. **(B)** progression-free patients at 6 months. *Abbreviations*: as in Figure S4.

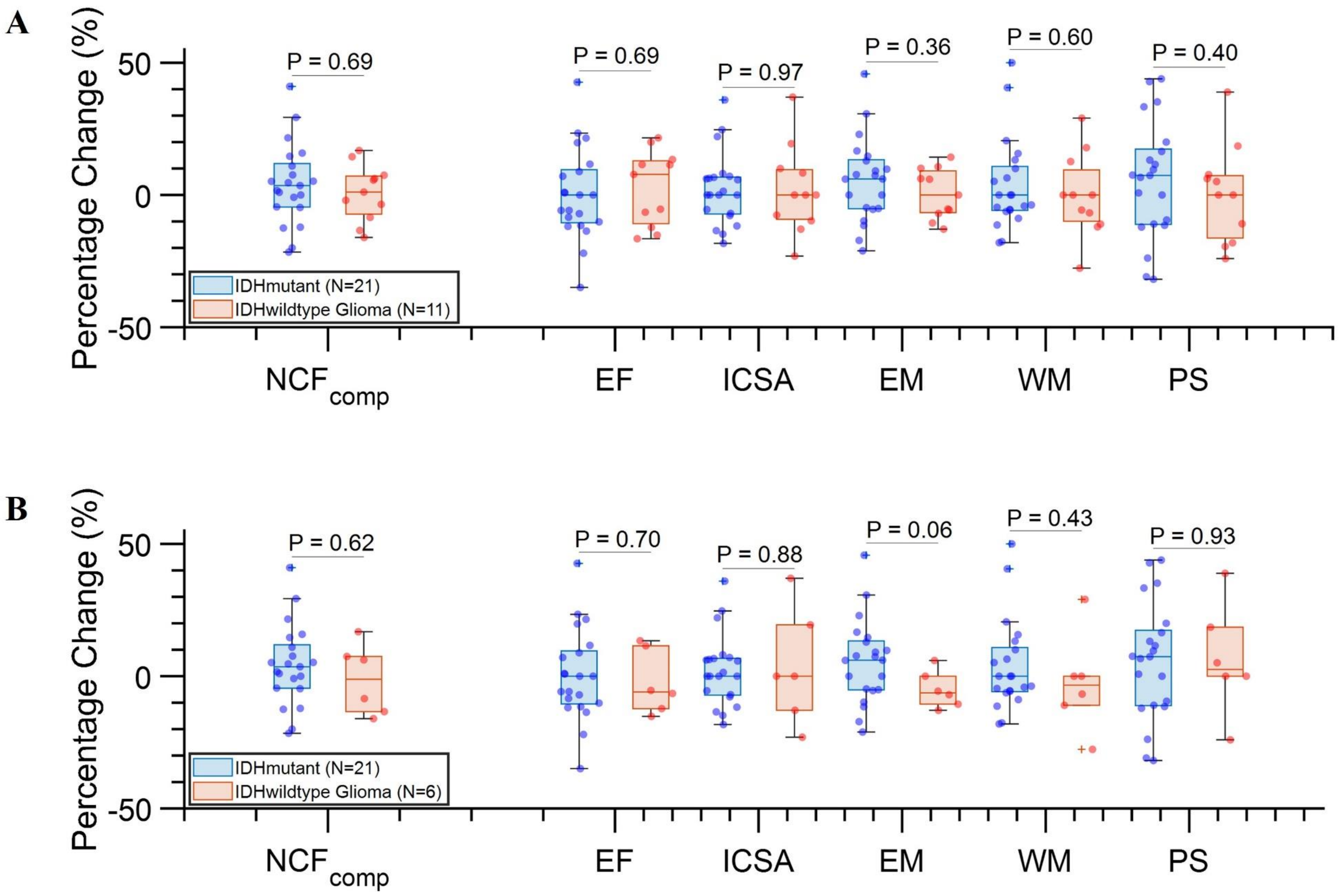

**Supplementary Figure S6: Patient-reported outcomes (PRO) vs. neurocognitive function (NCF) changes. (A-E)** Box plot showing the baseline, 6-month, and change between baseline and 6-month scores for MDASI-BT Symptom Severity, MDASI-BT Symptom Interference, MDASI-BT Cognitive Factor, Neuro-QOL, and LASA-QOL, respectively. **(F-J)** Scatter plot showing the percent change in fluid composite score from baseline to 6 months ($\Delta NCF_{Composite}$) versus the change in MDASI-BT Symptom Severity, MDASI-BT Symptom Interference, MDASI-BT Cognitive Factor, Neuro-QOL, and LASA, respectively. *Abbreviations*: MDASI-BT = The M.D. Anderson Symptom Inventory Brain Tumor Module; Neuro-QOL = the Neuro-Quality of Life Cognitive Function Short Form (Version2.0); LASA-QOL = the linear analogue self-assessment of self-perceived quality of life score.

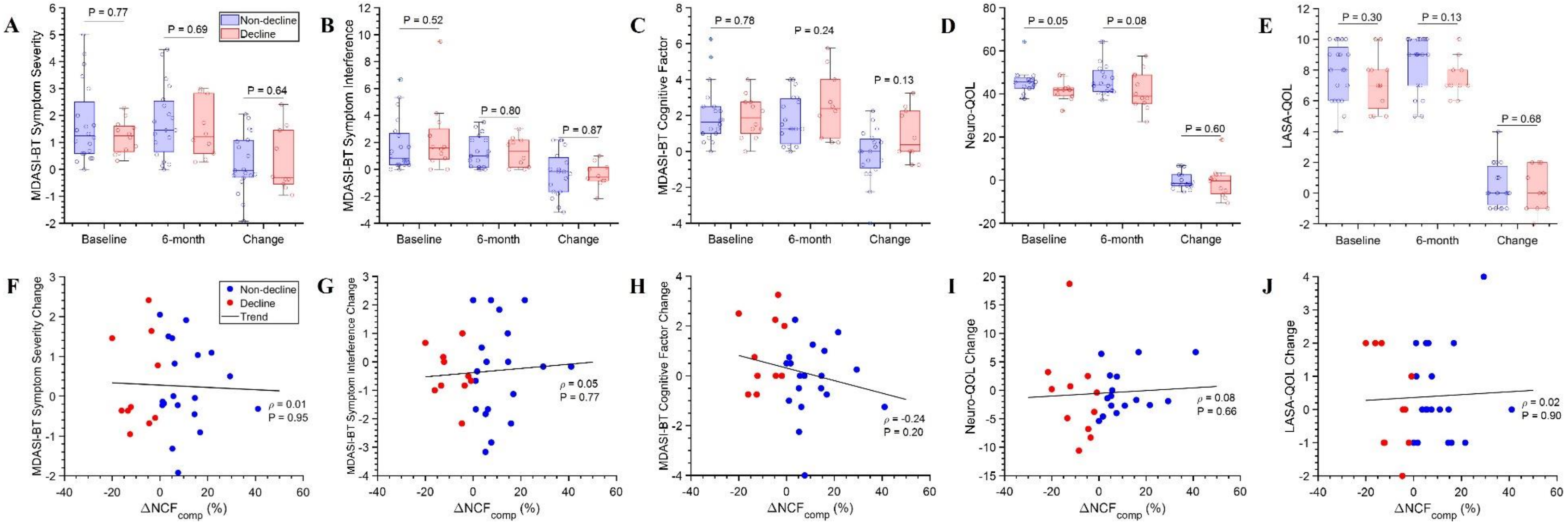

**Supplementary Figure S7: Contralateral functional connectivity (FC) matrices derived from ROIs in the hemisphere contralateral to the tumor.** **(A-C)** Composite FC matrix of the non-decline cohort at baseline, 6 months, and 6 months minus baseline. **(D-F)** Composite FC matrices for the decline cohort at baseline, 6 months, and 6 months minus baseline. Arrows indicate blocks with the strongest predictive performance for the change of NCF from the connectivity-regression analysis.

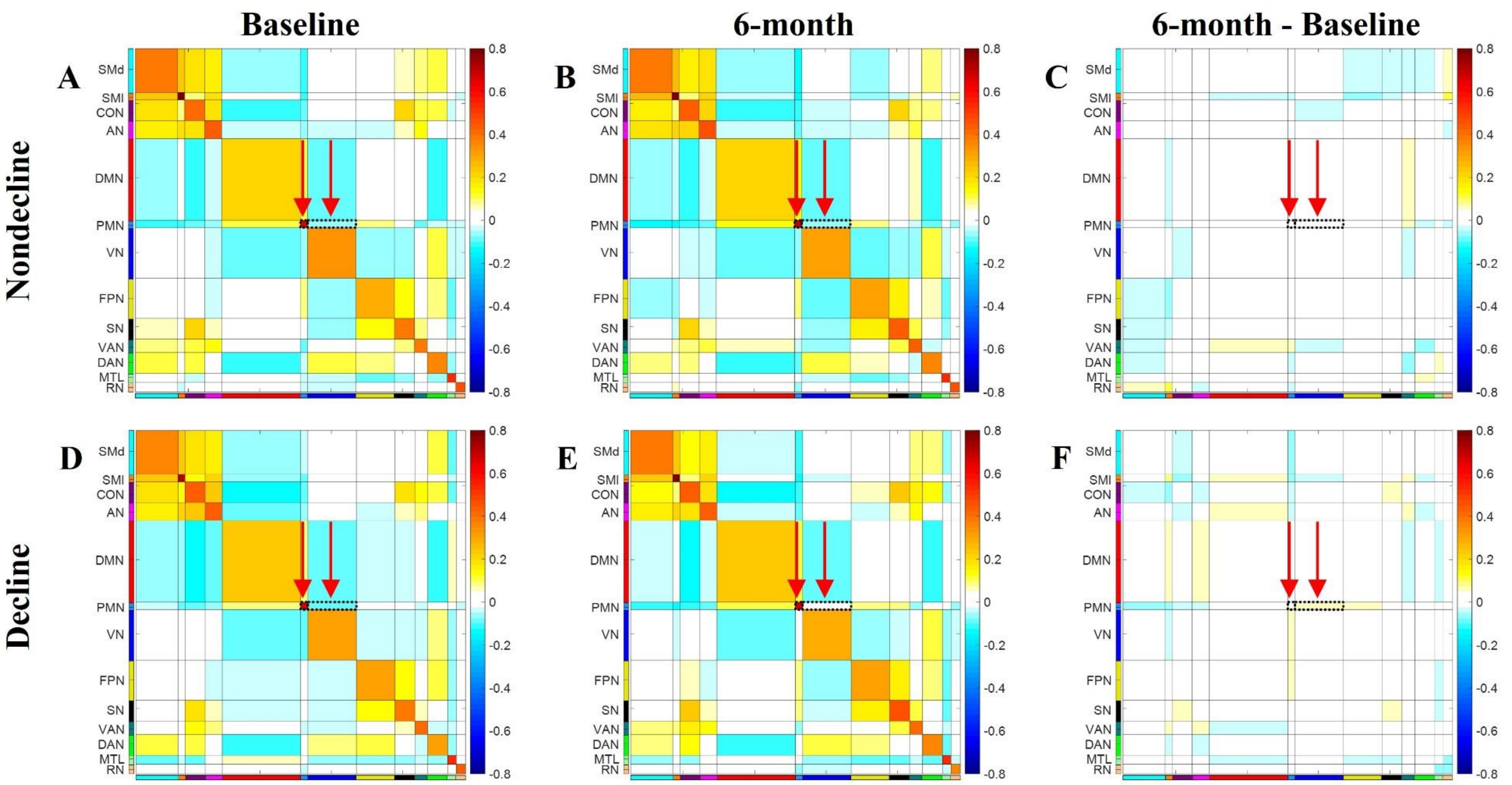

**Supplementary Table S1: Clinical and treatment characteristics of all patients enrolled.**

| | All (n=43) | Evaluable (n=32) | Non-evaluable (n=11) |
|---|---|---|---|
| Age | 49 (21-68) | 43 (21-68) | 56 (35-67) |
| Sex | | | |
| Male | 23 (54%) | 16 (50%) | 7 (64%) |
| Female | 20 (46%) | 16 (50%) | 4 (36%) |
| Race | | | |
| White | 36 (84%) | 28 (88%) | 8 (73%) |
| Black | 5 (12%) | 3 (9%) | 2 (18%) |
| Other | 2 (4%) | 1 (3%) | 1 (9%) |
| Hispanic Ethnicity | 3 (7%) | 3 (9%) | -- |
| KPS | | | |
| 70-80 | 10 (23%) | 5 (16%) | 5 (45%) |
| 90-100 | 33 (77%) | 27 (84%) | 6 (55%) |
| Highest Education | | | |
| High School | 14 (33%) | 10 (31%) | 4 (36%) |
| College | 22 (51%) | 17 (53%) | 5 (46%) |
| Graduate School | 7 (16%) | 5 (16%) | 2 (18%) |
| Tumor Type | | | |
| IDH-Mutant | 26 (60%) | 21 (66%) | 5 (45%) |
| IDH-Wildtype | 17 (40%) | 11 (34%) | 6 (55%) |
| EOR | | | |
| GTR | 19 (44%) | 12 (38%) | 7 (64%) |
| STR | 14 (32%) | 11 (34%) | 3 (27%) |
| Biopsy | 2 (5%) | 2 (6%) | -- |
| LITT | 6 (14%) | 6 (19%) | -- |
| No surgery (recurrence) | 2 (5%) | 1 (3%) | 1 (9%) |
| Tumor Sidedness | | | |
| Right | 29 (68%) | 20 (63%) | 9 (82%) |
| Left | 13 (30%) | 12 (37%) | 1 (9%) |
| Bilateral | 1 (2%) | -- | 1 (9%) |
| Tumor Grade | | | |
| 2 | 10 (23%) | 9 (28%) | 1 (9%) |
| 3 | 11 (26%) | 8 (25%) | 3 (27%) |
| 4 | 22 (51%) | 15 (47%) | 7 (64%) |
| IDH-Mutant Histology | | | |
| Astrocytoma | 15 (58%) | 12 (57%) | 3 (60%) |
| Oligodendroglioma | 11 (42%) | 9 (43%) | 2 (40%) |
| IDH-Wildtype MGMT Status | | | |
| Methylated | 10 (59%) | 7 (64%) | 3 (50%) |
| Unmethylated | 7 (41%) | 4 (36%) | 3 (50%) |
| RT Dose (Gy) | 60 (54-60) | 59.7 (54-60) | 60 (54-60) |
| RT Modality | | | |
| IMRT | 38 (88%) | 27 (84%) | 11 (100%) |

| | | | |
|---|---|---|---|
| PBT | 5 (12%) | 5 (16%) | -- |
| Concurrent TMZ | 28 (65%) | 21 (66%) | 7 (64%) |
| Adjuvant TMZ | 35 (81%) | 29 (91%) | 6 (55%) |

Abbreviations as in Table 1.

**Supplementary Table S2: Comparison of cognitive test scores between the decline vs non-decline cohorts.**

| | Non-decline (n=20) | Decline (n=12) | *P*-value |
|---|---|---|---|
| Baseline Fluid Composite | 84 (75-99) | 93 (82-105) | 0.36 |
| Baseline Executive Function Score | 91 (84-103) | 101 (84-108) | 0.40 |
| Baseline Attention Score | 77 (72-89) | 88 (70-93) | 0.32 |
| Baseline Episodic Memory Score | 98 (92-103) | 102 (91-108) | 0.42 |
| Baseline Working Memory Score | 96 (89-114) | 100 (78-125) | 0.85 |
| Baseline Processing Speed | 88 (70-95) | 94 (79-101) | 0.47 |
| 6-month Fluid Composite | 94 (85-109) | 84 (68-100) | 0.06 |
| 6-month Executive Function Score | 97 (88-109) | 89 (77-97) | 0.06 |
| 6-month Attention Score | 85 (74-94) | 79 (70-85) | 0.20 |
| 6-month Episodic Memory Score | 105 (92-115) | 95 (81-116) | 0.28 |
| 6-month Working Memory Score | 100 (90-117) | 100 (84-112) | 0.48 |
| 6-month Processing Speed | 94 (78-108) | 85 (61-92) | 0.07 |
| Percent Change of Fluid Composite [Median (IQR)] | 6.8 (3.8 – 15.5) | -10.3 (-15.4 – -3.8) | -- |
| Percent Change of Executive Function Score | 8.3 (-5.7 – 19.9) | -7.7 (-14.5 – -1.2) | 0.006 |
| Percent Change of Attention Score | 6.2 (0 – 9.6) | -8.7 (-14.5 – -1.4) | <0.001 |
| Percent Change of Episodic Memory Score | 6.1 (-4.0 – 12.3) | -5.0 (-12.3 – 9.8) | 0.10 |
| Percent Change of Working Memory Score | 0 (-5.6 – 13.1) | -4.9 (-10.7 – 0) | 0.21 |
| Percent Change of Processing Speed | 7.6 (0.2 – 30.0) | -11.2 (-24.0 – 4.6) | 0.005 |

**Supplementary Table S3: Comparison of patient-reported outcomes between the decline vs non-decline cohorts.**

| | Non-Decline (n=19) | Decline (n=10) | *P*-value |
|---|---|---|---|
| Baseline MDASI-BT Symptom Severity Score | [n=20]<br><br>1.3 (0.6-2.8) | [n=12]<br><br>1.3 (0.6 – 1.6) | 0.76 |
| Baseline MDASI-BT Symptom Interference Score | [n=20]<br><br>0.8 (0.3 – 2.8) | [n=12]<br><br>1.6 (0.7 – 3.3) | 0.51 |
| Baseline MDASI-BT Cognitive Factor Score | [n=20]<br><br>1.6 (1.0 – 2.5) | [n=12]<br><br>1.9 (0.9 – 2.8) | 0.77 |
| Baseline MDASI-BT Affective Factor Score | [n=20]<br><br>1.2 (0.7 – 4.2) | [n=12]<br><br>1.7 (0.8 – 2.5) | 0.83 |
| Baseline LASA QOL Score | [n=20]<br><br>8.0 (6.0 – 9.8) | [n=12]<br><br>7 (5.3 – 8.0) | 0.29 |
| Baseline Neuro-QOL Score | [n=16]<br><br>45.5 (42.7 – 48.1) | [n=11]<br><br>41.9 (38.9 – 42.9) | 0.051 |
| 6-month MDASI-BT Symptom Severity Score | 1.5 (0.5 – 2.6) | 1.2 (0.5 – 2.8) | 0.68 |
| 6-month MDASI-BT Symptom Interference Score | 1.0 (0.2 – 2.5) | 1.3 (0.1 – 2.2) | 0.78 |
| 6-month MDASI-BT Cognitive Factor Score | 1.3 (0.3 – 3.0) | 2.4 (0.7 – 4.3) | 0.23 |
| 6-month MDASI-BT Affective Factor Score | 1.8 (1.0 – 4.6) | 2.1 (0.7 – 3.7) | 0.55 |
| 6-month LASA QOL Score | 9.0 (7.0 – 10.0) | 7.0 (6.8 – 8.3) | 0.13 |
| 6-month Neuro-QOL Score | (n=20)<br><br>44.3 (41.1 – 51.4) | (n=12)<br><br>39.1 (35.5 – 49.0) | 0.08 |
| Change of MDASI-BT Symptom Severity Score | -0.1 (-0.3 – 1.1) | -0.3 (-0.6 – 1.5) | 0.63 |
| Change of MDASI-BT Symptom Interference Score | -0.2 (-1.7 – 1.0) | -0.6 (-0.9 – 0.3) | 0.91 |

| | | | |
|---|---|---|---|
| Change of MDASI-BT Cognitive Factor Score | 0 (-1 – 0.8) | 0.4 (-0.2 – 2.3) | 0.12 |
| Change of MDASI-BT Affective Factor Score | 0 (-0.6 – 1.6) | -0.3 (-1.3 – 1.5) | 0.54 |
| Change of LASA QOL Score | 0 (-1 – 2)) | 0 (-1 – 2) | 0.65 |
| Change of Neuro-QOL Score | [n=16]<br><br>-1.6 (-2.7 – 2.6) | [n=11]<br>-0.4 (-6.8 – 2.5) | 0.59 |

**Supplementary Table S4: Comparison of ipsilateral and contralateral FC matrices across individual patients.**

| Patient ID | Baseline (*P*-value) | 6-month (*P*-value) |
|---|---|---|
| 1. 1169_002 | 0.21 | **0.002** |
| 2. 1169_005 | 0.09 | 0.42 |
| 3. 1169_006 | 0.08 | 0.09 |
| 4. 1169_008 | 0.21 | 0.17 |
| 5. 1169_009 | 0.85 | **< 0.001** |
| 6. 1169_010 | 0.96 | 0.15 |
| 7. 1169_011 | 0.57 | 0.97 |
| 8. 1169_012 | 0.38 | 0.26 |
| 9. 1169_014 | 0.06 | **0.03** |
| 10. 1169_015 | 0.48 | 0.11 |
| 11. 1169_016 | 0.08 | **0.04** |
| 12. 1169_020 | 0.69 | 0.15 |
| 13. 1169_021 | 0.56 | **0.04** |
| 14. 1169_022 | 0.31 | 0.70 |
| 15. 1169_024 | 0.69 | 0.28 |
| 16. 1169_025 | **0.03** | 0.24 |
| 17. 1169_026 | 0.59 | 0.64 |
| 18. 1169_027 | 0.26 | 0.86 |
| 19. 1169_033 | **0.02** | **0.001** |
| 20. 1169_035 | 0.94 | 0.12 |
| 21. 1169_036 | 0.30 | 0.88 |
| 22. 1169_037 | 0.22 | **0.03** |
| 23. 1169_038 | 0.20 | 0.22 |
| 24. 1169_039 | 0.26 | **0.01** |
| 25. 1169_040 | 0.33 | **0.04** |
| 26. 1169_042 | 0.16 | **< 0.001** |
| 27. 1169_043 | 0.35 | 0.64 |
| 28. 1169_044 | 0.14 | 0.91 |
| 29. 1169_046 | 0.26 | **0.01** |
| 30. 1169_047 | **0.01** | 0.28 |
| 31. 1169_048 | 0.93 | 0.54 |
| 32. 1169_050 | 0.60 | 0.54 |
